# Relationships among Sleep Quality, Postural Alignment, Muscle Strength, and Psychological Health in Healthy Adults

**DOI:** 10.1101/2025.05.03.25326851

**Authors:** Jun-hee Kim, Sil-ah Choi, Sung-hoon Jung

## Abstract

**Background:** Sleep significantly affects physical and mental health, and influences factors such as posture, muscle strength, and psychological well-being. Despite its importance, modern lifestyle factors contribute to widespread sleep disturbances, highlighting the necessity for deeper insight into how sleep interacts with health indicators.

**Purpose:** This study aimed to investigate the complex relationships among sleep duration and quality, physical health indicators, and mental health status in healthy adults.

**Study design:** Cross-sectional study.

**Methods:** A total of 66 healthy adults aged between 20 and 60 years were assessed for sleep quality, depressive symptoms, sleep duration, spinal curvature, and thoracic muscle strength. Pearson’s correlation coefficient was used to analyze the relationships among these variables.

**Results:** Sleep duration was negatively correlated with the PSQI score (r = –0.344, p = 0.005) and kyphosis angle (r = –0.407, p = 0.001). Poor sleep quality positively correlated with depressive symptoms (r = 0.311, p = 0.011). The kyphosis angle was positively correlated with the thoracic extensor muscle strength (r = 0.675, p < 0.001).

**Conclusions:** This study demonstrates significant associations among sleep quality, spinal alignment, and mental health. This emphasizes the need for integrated clinical approaches that address sleep, posture, and psychological well-being to enhance overall health outcomes.

## INTRODUCTION

Sleep is a vital component of human health that influences numerous physiological and psychological processes. Adequate sleep is crucial for optimal cognitive functioning, emotional regulation, and physical health.^1^ Despite their importance, sleep disorders and inadequate sleep duration are prevalent and pose significant risks to the overall well-being.^2^ In modern society, lifestyle changes such as increased screen time, work-related stress, and irregular schedules have contributed to widespread sleep insufficiency, making this issue even more relevant.^3,4^ Poor sleep quality and short sleep duration have been linked to numerous adverse health outcomes such as an increased risk of obesity, diabetes, cardiovascular disease, and mental health disorders.^5,6^ Emerging evidence also suggests that poor sleep may impair neuromuscular recovery, increase systemic inflammation, and alter the autonomic nervous system balance, further compounding its impact on health.^7–9^

Physical health is also closely associated with sleep quality.^10^ Kyphosis, the forward curvature of the spine commonly observed in older adults, can significantly affect mobility and quality of life.^11^ The degree of kyphosis can influence an individual’s ability to perform daily activities, potentially leading to increased dependency and reduced physical function.^12^ Muscle strength is another critical indicator of physical health that reflects an individual’s ability to maintain functional independence.^13^ Poor muscle strength is associated with a high risk of falls, fractures, and disabilities.^14,15^ Recent studies have highlighted that disrupted sleep may contribute to sarcopenia and muscle weakness, suggesting a direct link between sleep disturbances and a decline in musculoskeletal health.^16^ Mental health, which encompasses conditions such as depression and anxiety, is profoundly influenced by sleep.^6^ Numerous studies have highlighted the bidirectional relationship between sleep disturbances and depression, where poor sleep can exacerbate depressive symptoms, and conversely, depression can lead to impaired sleep.^17,18^ Additionally, chronic sleep deprivation has been associated with heightened emotional reactivity, impaired stress-coping mechanisms, and alterations in neurotransmitter systems such as serotonin and dopamine, all of which may mediate the relationship between sleep and mood disorders.^19–21^

The relationships among sleep, physical health, and mental health are deeply interconnected and bidirectional.^17,18^ Sleep disturbances have been shown to negatively affect muscle strength, postural alignment, and emotional regulation, while physical dysfunction and mental health conditions such as depression and anxiety can lead to poor sleep quality, creating a self-reinforcing cycle of impairment.^11,14^ This vicious cycle may accelerate functional decline, increase dependency, and reduce overall wellbeing, particularly among older adults.^12,15^ In addition, poor sleep is linked to increased systemic inflammation, impaired neuromuscular recovery, and autonomic nervous system dysregulation, underscoring its widespread physiological impact.^7–9^

Despite the growing recognition of the interrelated nature of sleep quality, physical health, and mental health, few studies have concurrently investigated these domains within an integrated analytical framework. Previous research has predominantly examined each factor independently, constraining a comprehensive understanding of their dynamic interplay. Sleep disturbances may serve as contributing factors to declines in both physical function and psychological well-being, underscoring the necessity of simultaneous assessment.

Therefore, this study aimed to explore the complex relationships among sleep duration and quality, physical health indicators, and mental health. By analyzing these correlations, we aimed to contribute to the growing body of knowledge on how sleep influences overall health. This study investigated these relationships, focusing on sleep duration and quality, physical health indicators such as kyphosis angle and thoracic extensor muscle strength, and mental health. We hypothesized that sleep duration and quality would interact with physical and mental health such that poor sleep would exacerbate the negative relationship between postural deviations and psychological distress.

## METHODS

### Participants

This study recruited 66 healthy adults (29 males and 37 females) aged 20–60 years. The participants were recruited through community posts and university bulletin boards. The inclusion criteria required that participants have no history of musculoskeletal, neurological, or psychiatric disorders that could affect their sleep patterns, posture, or physical performance. Exclusion criteria included any diagnosed sleep disorders (e.g., insomnia, sleep apnea), recent musculoskeletal injuries (within the last six months), or ongoing pharmacological treatments affecting sleep or mood. The experimental procedure was approved by the Institutional Review Board (IRB) of Yonsei University (approval number: 1041849-202101-BM-004-02). Prior to participation, all participants were fully informed about the purpose and details of the study and voluntarily agreed to participate after providing written informed consent.

**Table 1.** Characteristics of the study participants

|  | Male (n = 29) | Female (n = 37) | Total (n = 66) |
| --- | --- | --- | --- |
| Age (years) | 32.55 ± 5.21 | 32.11 ± 7.97 | 32.30 ± 6.85 |
| Height (cm) | 174.51 ± 6.09 | 162.03 ± 6.37 | 167.52 ± 8.80 |
| Weight (kg) | 75.84 ± 11.55 | 63.05 ± 12.05 | 68.67 ± 13.37 |

### Procedure

The participants completed subjective questionnaires and underwent objective sleep and physical health assessments. Sleep and psychological assessments: Subjective sleep quality was measured using the Pittsburgh Sleep Quality Index (PSQI), and depressive symptoms were assessed using the Beck Depression Inventory (BDI).^22,23^ Sleep duration and patterns were objectively monitored using the Galaxy Fit2 (Samsung Electronics, South Korea), a wrist-worn actigraphy device worn continuously for seven consecutive days (Figure 1). The average values from the seven-day data were used for analysis. The kyphosis angle was measured in a natural standing posture using the Spinal Mouse® (Idiag AG, Fehraltorf, Switzerland). While standing upright with arms relaxed, the examiner guided the Spinal Mouse® device along the spinous processes from C7 to S3. The participants were instructed to stand comfortably in their habitual posture (Figure 2). Thoracic extension strength was measured in the quadruped position using a Smart KEMA Measurement System (KOREATECH Co., Ltd., Seoul, South Korea). The participants assumed a modified quadruped position with both knees flexed to 90° and the trunk placed over a cushioned bench. The pelvis and lower legs were stabilized using strips. The upper body was positioned forward and the participants clasped their hands behind their heads while performing maximal isometric thoracic extension against a resistance band connected to the Smart KEMA sensor system (Figure 3). The mean value of three trials, each involving a 5-s maximal contraction, was used for the analysis. For each trial, the middle 3 s of contraction was extracted and analyzed.

**Figure 1.**
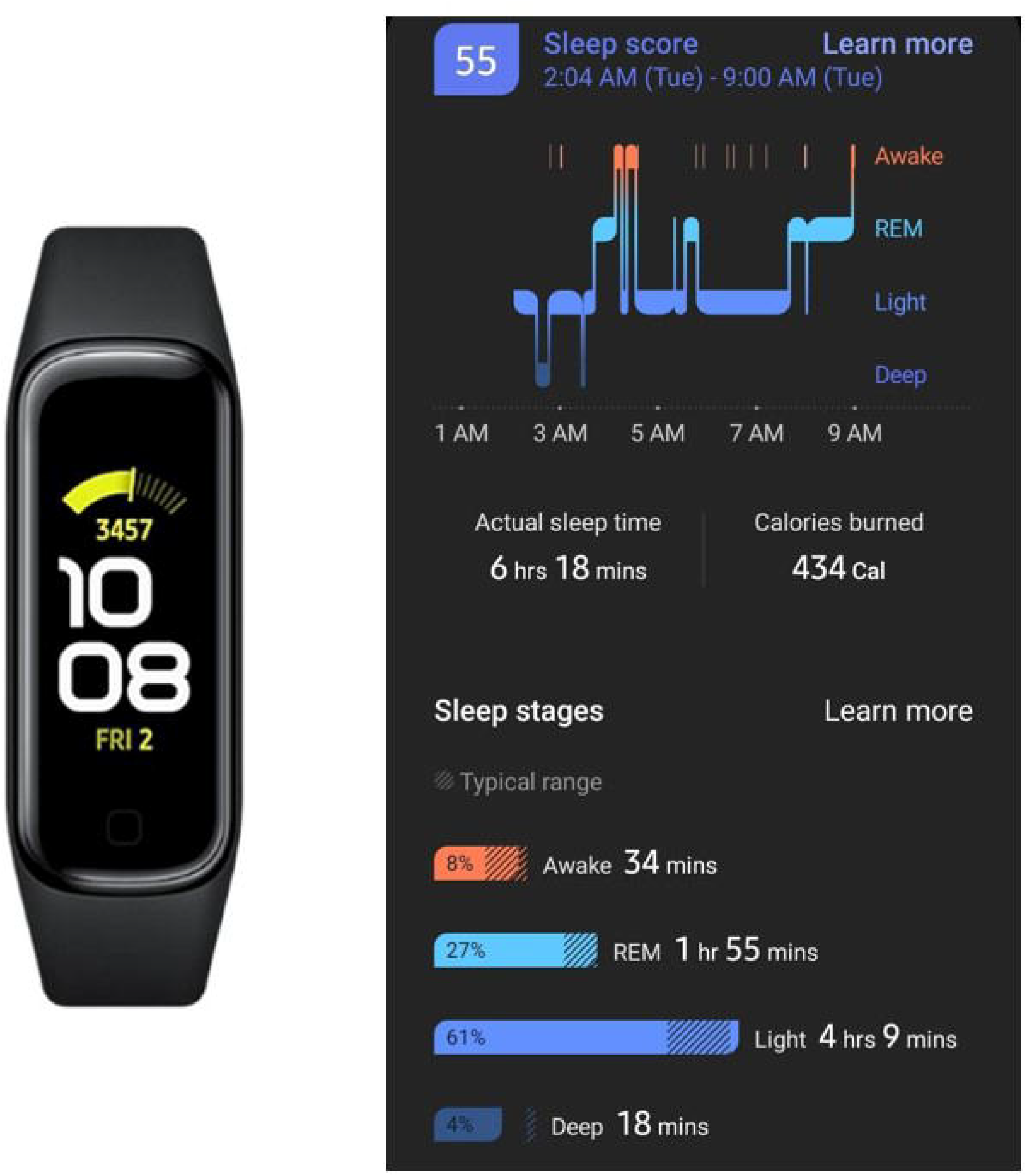
Sleep monitoring using Galaxy Fit2

**Figure 2.**
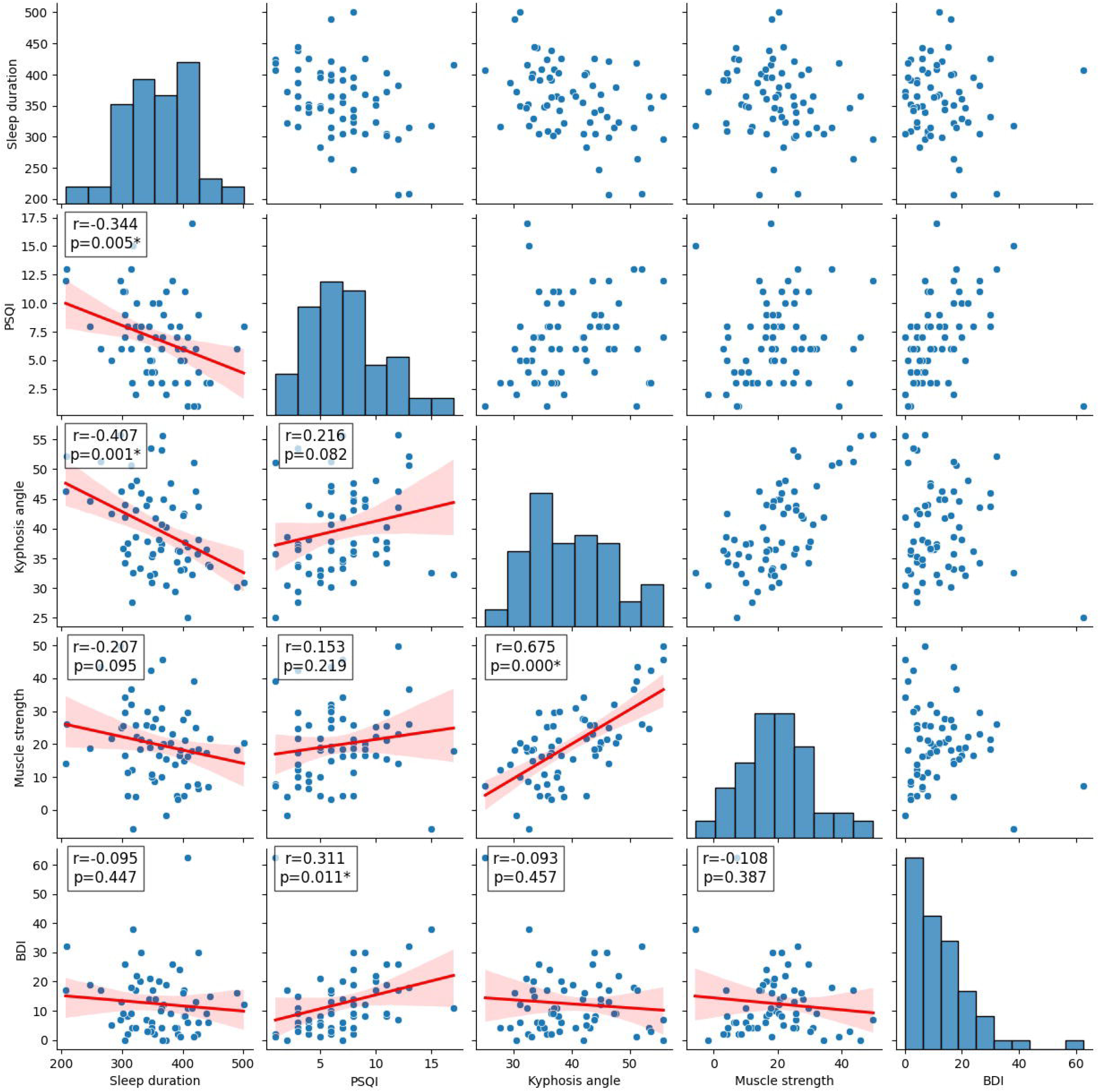
Kyphosis angle measurement using Spinal Mouse

**Figure 3.** Assessment of thoracic extensor strength in the quadruped position

### Statistical analysis

All analyses were performed using Python in the Google Colab environment (Google LLC, Mountain View, CA, USA). Descriptive statistics (mean ± standard deviation) were calculated for all variables. The Shapiro–Wilk test was used to verify the normality of continuous variables. Pearson’s correlation analyses were conducted to assess the relationships among sleep duration, PSQI scores, kyphosis angle, muscle strength, and BDI scores. Statistical significance was set at p < 0.05.

## RESULTS

Pearson’s correlation analysis was conducted to examine the relationships among sleep duration, PSQI, kyphosis angle, muscle strength, and BDI scores (Figure 4).

**Figure 4.** Correlation matrix among variables

Sleep duration was significantly negatively correlated with PSQI (r = –0.344, p = 0.005) and kyphosis angle (r = –0.407, p = 0.001), indicating that longer sleep duration was associated with better sleep quality and lower kyphosis angle. However, no significant correlations were observed between sleep duration and muscle strength (r = –0.207, p = 0.095) or BDI scores (r = –0.095, p = 0.447).

The PSQI was significantly positively correlated with BDI scores (r = 0.311, p = 0.011), suggesting that poor sleep quality was associated with higher depressive symptoms. No significant correlations were found between PSQI and kyphosis angle (r = 0.216, p = 0.082) or muscle strength (r = 0.153, p = 0.219).

The kyphosis angle showed a significant positive correlation with muscle strength (r = 0.675, p < 0.001), indicating that a greater kyphosis angle is associated with greater thoracic extensor muscle strength. However, no significant correlation was found between the kyphosis angle and BDI scores (r = –0.093, p = 0.457). Muscle strength was not significantly correlated with BDI scores (r = –0.108, p = 0.387).

**Table 2.** Descriptive statistics of variables

| Variable | Mean ± SD |
| --- | --- |
| Sleep duration (min) | 359.24 ± 57.53 |
| PSQI | 6.82 ± 3.49 |
| Kyphosis angle ($^{\circ}$) | 39.82 ± 7.26 |
| Muscle strength (kg) | 19.83 ± 11.24 |
| BDI | 12.36 ± 10.71 |

## DISCUSSION

This study investigated the relationship between sleep duration and quality and physical and psychological health in healthy adults. The key findings revealed that longer sleep duration was significantly associated with better subjective sleep quality (lower PSQI scores) and reduced kyphosis angles. Additionally, poor sleep quality has been linked to greater depressive symptoms, and an increased kyphosis angle has been associated with reduced thoracic extensor muscle strength.

The observed negative correlation between sleep duration and kyphosis angle suggests that sufficient sleep may contribute to the maintenance of spinal alignment and postural stability. This aligns with the findings of a previous study showing that sleep facilitates muscle recovery and may prevent cumulative musculoskeletal strain, which could otherwise result in postural deviations, such as hyperkyphosis.^7^ Previous studies have shown that sleep deprivation negatively affects postural control by impairing proprioception and neuromuscular coordination.^24^ Similarly, inadequate sleep has been shown to increase the risk of postural instability and alter gait mechanics, which may contribute to long-term changes in the spinal curvature.^25–27^

The significant relationship between the PSQI and BDI scores highlights the critical role of sleep quality in mental health. Given that poor sleep can exacerbate emotional dysregulation and depressive symptoms, this result supports prior literature that advocates sleep-focused interventions as a part of comprehensive mental health care. Consistent with our findings, Johansson et al. (2021) demonstrated that individuals with chronic insomnia have a significantly higher risk of developing depression.^28^ Furthermore, a meta-analysis by Fang et al. (2019) confirmed that treating sleep disturbances can reduce depressive symptoms and support sleep as a modifiable factor in mental health interventions.^29^

This study found a positive correlation between kyphosis angle and thoracic extensor muscle strength in a non-older population, which contrasts with the findings typically seen in older adults. Rather than reflecting degeneration, this relationship may indicate a compensatory mechanism, in which individuals with increased kyphosis involuntarily recruit trunk muscles more intensely to maintain balance and posture. Prior research has shown that individuals with postural changes or low back pain exhibit heightened trunk muscle activation, even without targeted training, likely as a protective or stabilizing response.^30,31^ These findings suggest that increased muscle strength may not result from formal exercise but from neuromuscular adaptation to an altered posture.

The findings of this study emphasize the need for integrated health interventions that concurrently address sleep disturbance, postural health, and psychological well-being. Clinically, this highlights the importance of routine assessments of sleep patterns and postural alignment as part of comprehensive health evaluations, even in non-older and otherwise healthy adults. Promoting adequate sleep duration and improving sleep quality may not only enhance mental health outcomes such as reducing depressive symptoms, but may also support musculoskeletal function and prevent postural deterioration. Furthermore, recognizing the compensatory muscular adaptations to postural deviation offers valuable insights for clinicians when designing targeted rehabilitation or preventive exercise programs. These insights underscore the necessity for a holistic and multidisciplinary approach to clinical practice that aims to promote long-term physical and psychological health.

This study had certain limitations. First, the cross-sectional design limited the ability to determine causal relationships among sleep, physical health, and psychological health. Second, the study sample consisted of only healthy adults aged 20–60 years, which may restrict the generalization to older adults or clinical populations with musculoskeletal or mental health conditions. Additionally, while sleep was measured objectively using actigraphy, this method may not fully capture the detailed sleep architecture compared to polysomnography. Physical assessments were limited to kyphosis angle and thoracic extensor muscle strength, excluding other relevant factors such as balance or core stability. Future research should adopt longitudinal or interventional designs to clarify causal pathways and include a broader population. The incorporation of more comprehensive physical assessments and advanced sleep evaluation tools could further enhance our understanding of the interplay among sleep, posture, muscle strength, and mental health.

## CONCLUSIONS

This study demonstrated that longer sleep duration was significantly associated with better subjective sleep quality and reduced kyphosis angle, highlighting the potential role of adequate sleep in maintaining spinal alignment and posture. Furthermore, poor sleep quality was positively correlated with depressive symptoms, emphasizing the critical connection between sleep and mental health. These findings underscore the importance of integrating sleep management with strategies to promote musculoskeletal health and psychological well-being.

## Data Availability

All data produced in the present study are available upon reasonable request to the authors

## ACKNOWLEDGEMENTS

This research was supported by Basic Science Research Program through the National Research Foundation of Korea (NRF) funded by the Ministry of Education (grant number: NRF- 2020R1A6A3A01099963)

## STATEMENT

The authors take full responsibility for the data, the analyses and interpretation, the conduct of the research, full access to all the data, and the right to publish any data.

## DECLARATION

All authors and contributors agree to the conditions outlined in the Authorship and Contributorship section of the Information for Authors.

## ETHICAL APPROVAL

The present study was approved by the Institutional Review Board of Yonsei University (Seoul, Korea) (approval no. 1041849-202101-BM-004-02).

